# Co-Designing Pay-It-Forward Strategies to Improve Retention in Cervical Cancer Care in Kenya: A Formative Participatory Study

**DOI:** 10.64898/2026.09.10.26362751

**Authors:** Harriet Fridah Adhiambo, Anne Trolard, Dorothy Mangale, Eric Thuo, Philippa Kadama Makanga, Lucy Akoo, Jerome Katumba, Phiona Adagi, Betsy Abente, Beryne Odeny, Elizabeth A. Bukusi, Byron Powell, Bettina Drake, Dickens Onyango, Elvin Geng, Joseph D. Tucker, Thomas Odeny

**Author notes:** **Corresponding Author:** Harriet Fridah Adhiambo /.

## Abstract

**Background:** Retention is the bridge between diagnosis and survival in low and middle-income countries, including Kenya, where the cost of cervical cancer treatment can amount to several years of an average woman’s earnings. Pay-it-forward (PIF), a prosocial approach in which an individual receives a gift and then considers supporting another person, offers a community-driven strategy to reduce barriers and improve engagement and retention in cancer care. This study explored opportunities to integrate PIF strategies into cervical cancer care through a human-centered design (HCD) workshop.

**Methods:** We conducted a two-day HCD workshop in Kenya, informed by the WHO/UNICEF normative guide on co-creation with women with cervical cancer (on treatment or survivors), community representatives, and oncology care providers. Experience diagramming mapped the patient journey and identified barriers to retention. A designer-led team then developed the initial PIF prototypes, which were iteratively co-designed and refined with participants to align with their needs and preferences. Audio recordings and photographs of the co-design activities were transcribed and analyzed using rapid, team-based qualitative synthesis.

**Results:** Among 25 participants, 52% (n=13) were women with cervical cancer and 48% (n=12) were health care providers or community representatives. Participants identified costs, extenuating circumstances, treatment side effects, health system challenges, discrimination, fear, and anxiety as barriers to retention. These barriers informed the co-design of six PIF component strategies: peer navigation, transport voucher fund, integrated service gift, a message board in clinic waiting areas, health insurance support fund, and a basket of kindness at checkout. Three of these (peer navigation, transport voucher fund, and health insurance support) have been effectively implemented in Kenya before for other health services. Participants noted that including both monetary and non-monetary nudges was essential but emphasized the need for transparency and oversight in monetary approaches.

**Conclusion:** The Kenyan context provided rich opportunities for PIF component strategies to explore and enhance retention in cancer care. We hypothesize that PIF may improve retention by activating social capital through reciprocity, empathy, and a sense of belonging. Future work should pilot these PIF strategies to assess feasibility, acceptability, and appropriateness, and determine whether reciprocal giving can be sustained.

## Introduction

The burden of cervical cancer continues to rise, especially in low-and middle-income countries (LMICs), which contribute to nearly 90% of the global cervical cancer burden.^1,2^ In 2024, cervical cancer was estimated to cause approximately 604,000 new cases and 280,000 deaths globally, with the vast majority occurring in LMICs.^1^ Substantial investments have been made in cervical cancer prevention, screening, and early detection programs, but these efforts have not yet been fully realized, as many women in LMIC continue to be diagnosed with advanced disease.^3,4^ The gap between these investments and health outcomes reflects persistent implementation challenges across the cervical cancer care continuum. For women diagnosed with cervical cancer, sustained engagement after diagnosis is critical for treatment initiation, completion, survivorship, and access to palliative care.^5^ Across Africa, an estimated 15-82% of women are lost to follow-up (LFTU) after diagnosis, impacting overall cancer management..^6–8^

Retention challenges in the cervical cancer care continuum are multifaceted but primarily driven by financial constraints, particularly non-medical expenses such as accommodation and transport, as well as limited access to comprehensive cancer care services.^9,10^ Non-financial factors, including psychosocial distress, fear and stigma, treatment-related challenges, limited social support, and difficulties navigating health systems, also contribute to treatment delays and loss to follow-up.^11^ If these challenges are not addressed, women will continue to experience treatment delays, avoidable morbidity and mortality, and increased burden on already constrained health systems.^12^

“Harambee,” a Kiswahili word meaning “pulling together” describes a practice in which communities pool resources to support members in need. For example, this includes raising funds for funerals, school fees, and the construction of schools and health facilities. This culture of generosity and shared responsibility is deeply established in Kenya. Pay-it-forward (PIF) is a prosocial intervention in which an individual receives a gift and then is invited to donate toward the care of a future recipient. Conceptually, PIF aligns closely with the spirit of Harambee, as it emphasizes mutual aid and collective responsibility. Randomized and quasi-experimental trials in Asia have shown that PIF increases uptake of human papillomavirus (HPV), hepatitis, and influenza vaccines, as well as testing for sexually transmitted infections, with voluntary donations covering up to 40% of service costs.^13–17^ That evidence, however, comes from preventive, low-cost services. Whether PIF can be adapted to a longitudinal, high-cost illness and whether it can align with an indigenous reciprocity norm, such as Harambee, is unknown. A human-centered design (HCD) approach to co-design PIF strategies to improve retention in cervical cancer care in western Kenya was undertaken, as formative work preceding feasibility testing.

## Methods

### Study setting

This study was conducted at the Kenya Medical Research Institute (KEMRI), which is co-located within the Jaramogi Oginga Odinga Teaching and Referral Hospital (JOOTRH) in Kisumu, Kenya. JOOTRH is a level VI facility and a major referral center serving a catchment population of approximately 10 million people across Western Kenya.^18^ It offers screening, diagnostic evaluation, surgery, chemotherapy, and palliative care. It does not offer radiation therapy, and patients who need radiotherapy travel to other regions. Cancer services are financed through out-of-pocket payment, the mandatory national health insurance scheme (Social Health Authority or SHA), or private insurance. Although SHA covers major components of cervical cancer care, including imaging, chemotherapy, radiotherapy, and brachytherapy, patients still incur costs not fully covered by insurance, including transportation, accommodation, caregiver expenses, and lost income.^19^ The cost of cancer care is substantial relative to household income, ranging from approximately US<$>1\comma \; 328 to more than US<$>9,663 depending on the disease stage and treatment requirements. The average income for a Kenyan woman is approximatelyUS<$>1\comma \; 128/year\comma \; while SHA provides oncology benefits of up to US<$>6,185 per patient, an equivalent of more than five years of the average earnings.^20,21^ These costs are particularly burdensome for lower-quantile-income women and may result in catastrophic situations.

## Study design and rationale

We aimed to learn directly from cervical cancer patients how a PIF intervention might support them by identifying the exact points in their cancer journey where resources are needed. We hosted a two-day HCD workshop in July 2025 with two goals: 1) mapping the patient journey in cervical cancer care to identify key pain points (Day 1), and 2) developing, iterating, and refining several PIF prototypes matched to those pain points for subsequent testing (Day 2). An HCD approach was appropriate because it centers the experiences of end users to elicit preferences and needs, increasing the likelihood that a) problem statements will reflect lived realities and b) the resulting strategies will be matched to patients’ needs and preferences. Design activities were guided by the double-diamond framework^22^ (Figure 1) and the WHO/UNICEF normative guide on co-creation.^23,24^ The process begins by discovering and defining the problem (first diamond), followed by iteratively developing solutions through brainstorming and testing to deliver a product that optimizes the fit between the design object and the end user (second diamond). Reporting of this work follows the recommendations of Bazzano et al. for comprehensive, systematic, and transparent documentation of human-centered design processes.^25^

**Figure 1:**
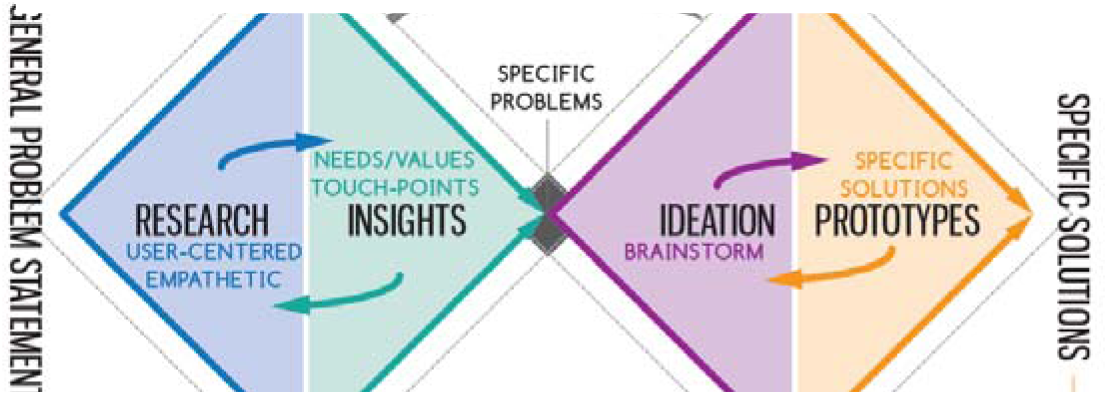
The Double Diamond Design Process

## Participants

### Design and Facilitation Team

The workshop was led by a multidisciplinary team comprising a design consultant (AT), a gynaecologic oncologist (JK), a palliative care nurse (LA), and two study coordinators (HFA and PKM).

A month prior to the workshop, the design consultant led three structured 1.5-hour training sessions for the study team by videoconference, covering design orientation to the study, overview and principles of HCD, a review of the double-diamond framework, and hands-on practice with HCD tools, including experience diagramming and affinity clustering.

We used purposive sampling to ensure that participants had direct experience of or a role in supporting cervical cancer care. Eligibility criteria were women diagnosed with cervical cancer and are currently on treatment or survivors, women diagnosed with cervical cancer and also living with HIV, health care providers at oncology clinics (nurses, oncologists, and clinical officers), and community representatives. 13 women were identified through clinic records and referrals from providers, whereas community advocates and health providers were identified in consultation with oncology clinic leadership. The team invited participants by phone call. Of 26 potential participants approached, 25 agreed to join, and one provider declined due to personal commitments. See Table 1 for a summary of participant characteristics.

**Table 1:** Participant Characteristics

| Patients with Cervical Cancer (n=13) |  |  |
| --- | --- | --- |
|  | Female (n) | % |
| Age |  |  |
| 40-50 | 8 | 62% |
| 51-60 | 3 | 23% |
| >60 | 2 | 15% |
| HIV Status |  |  |
| Living with HIV | 10 | 77% |
| Not Living with HIV | 3 | 13% |
| **Health Providers and Community Representatives (n=12)** |  |  |
|  | **Male (n, %)** | **Female (n, %)** |
| Cadres |  |  |
| Nurses | 1 (17%) | 5 (83%) |
| Clinical Officers | 1 (50%) | 1 (50%) |
| Oncologists | 1 (100%) | 0 |
| Community Representatives | 2 (50%) | 1 (50%) |

## Procedures and data collection

Data collection and design activities were organized according to the four phases of the Double Diamond framework: discover, define, develop, and deliver. The two-day workshop focused on the discover, define, and develop phases. The delivery phase, which will involve testing and implementing the co-designed PIF strategies, was beyond the scope of this formative study. Data collection throughout the workshop involved detailed notetaking and audio recording of discussions and activities. Workshop activities were conducted in English, Kiswahili, and Dholuo.

## Day 1: Discover & Define

Prior to the workshop, the study team reviewed literature on barriers to retention in cervical cancer treatment after diagnosis and identified the following touchpoints: preparing for the visit, traveling to the clinic, registering and waiting, clinical examination, laboratory and pharmacy visits, checking out, and preparing for the next visit .^26,27^ On Day 1 of the workshop, participants were introduced to the PIF concept through a role-play exercise; and women with lived experience of cervical cancer shared the experiences of their cancer care journey, including the people, places, and things at each touchpoint, as well as what was positive and what was negative at each touchpoint. Participant contributions were documented in real time to create a shared-experience diagram that represents the patient journey.

Following experience diagramming, negative experiences identified across the experience diagram were clustered (thematically grouped) and ranked by intensity through group discussions to identify which problems presented the greatest opportunity for the PIF intervention. These priority areas formed the basis of PIF prototype development.

## Day 2: Develop

Between days 1 and 2, the design consultant led a team of four study facilitators to brainstorm and create initial prototypes of the PIF interventions using storyboarding templates. This marked the first iteration of the prototype development. Participants were presented with the initial prototypes in small groups and asked to critique them by answering three questions: 1) Can you imagine this happening at your clinic? If not, why? 2) Would this help patients? 3) Would you participate? The goal of this session was to qualitatively assess the feasibility and acceptability of each prototype. Groups reported out, and responses were audio-recorded and organized, forming the basis of prototype revisions. Following the critique, participants worked in small groups to redesign and refine prototypes. Using new storyboard templates, each group revised the proposed PIF strategies and presented the refinements in a plenary. These participant-led refinements resulted in a second and final set of co-designed PIF prototypes for subsequent testing.

## Data Analysis

The primary outputs of the audio-recording analysis included 1) problem definition and insights into barriers and facilitators of engagement; and 2) participant critiques and ideas for each prototype PIF intervention. The purpose of the analysis was to synthesize participants’ experiences, critiques, and recommendations across workshop activities and use these findings to inform iterative development of the PIF strategies. Findings from the analysis were integrated with the workshop outputs to identify priority modifications and refine prototypes into final PIF strategies. Audio recordings of the workshop activities, including experience diagramming, prototype critique, and iterative prototyping sessions, were transcribed verbatim. We used rapid qualitative analysis through an iterative, team-based synthesis approach. Rather than a qualitative analysis software, the team systematically reviewed, organized, summarized, and clustered data through structured discussions and consensus to identify themes, prioritize implementation challenges, and refine intervention prototypes. HAF, PO, and RKA reviewed the transcripts and facilitator notes to identify patterns in the data and participant responses to the proposed PIF strategies. Similar observations were grouped and compared across participants and activities.

## Team debriefing and reflexivity

Debrief sessions with the study investigator team, workshop facilitators, and design consultant were held at the end of each workshop day. The sessions involved reviewing emerging themes from each activity to assess alignment with participant input, comparing insights with existing literature, and interpreting new findings from the co-design process. We recognized that differences in professional roles, lived experience, and relationship with the health system could influence participant contributions and the research team’s interpretation of the findings. To promote reflexivity, facilitators elicited perspectives across stakeholder groups and daily debriefs were used to identify areas of agreement and divergence. When perspectives differed, women’s lived experiences were prioritized for patient-experience issues, while provider and community perspectives informed feasibility and implementation. Prototype refinements remained grounded in participants’ input.

## Results

### Participant characteristics

The design workshop included women with lived experience of cervical cancer, both on treatment and post-treatment, oncology providers, and community representatives. Most patient participants (77%) were living with HIV and receiving antiretroviral therapy. Table 1 below summarizes participant characteristics.

We present the process by which we developed PIF prototypes through a structured human-centered design approach.

### Day 1: Participant reactions to the role play

Participants engaged with the PIF role play by sharing reflections grounded in their lived experiences with cancer care. They described the PIF approach as capable of generating motivation, hope, positive feelings, and goodwill within the clinic, factors they viewed as important during the cancer care journey. Participants also anticipated that the intervention could support retention in care, noting that providers would be encouraged to see patients remain engaged in treatment. Receiving services through the PIF model (i.e., care supported by an anonymous donor) was described as a meaningful form of encouragement that could motivate patients to continue attending treatment sessions as scheduled.

*“We really need this kind of encouragement to continue with our care because we have other eventualities that crop up that make people default in their treatment.” (PID 004)*

### Day 1: Patient experiences across the cervical cancer care journey

Fifty-five pain points were identified across the seven clinic visit touchpoints. Experience diagramming revealed that although the women’s experiences of cervical cancer care varied across the clinic journey, their specific pain points were less unique to certain individuals but rather pitfalls that anyone could experience while navigating a complex system. They often began the journey motivated to receive care and treatment, but this motivation was repeatedly tested by financial, logistical, health-system, and treatment-related challenges at different point along the care journey.

#### a) Preparing for and getting to the clinic

Participants described preparing for the clinic visits by managing 1) personal, family, and household duties; 2) their daily medications; and 3) the logistics of visiting the clinic (e.g., checking appointment cards, arranging transportation, and potentially a caregiver if too weak to travel). Despite these competing priorities, they were motivated to arrange it all to seek care, meet with the providers, discuss their health concerns, and hopefully get better. There were events that could disrupt their intentions, including family emergencies (e.g., a child’s illness) and the costs of care and transport. Participants said they could sometimes borrow money from relatives or friends but deemed it largely unsustainable because treatment made it difficult to work and repay the borrowed funds.

#### b) Registration and waiting

Arrival at the clinic marked a shift in their ability to troubleshoot problems and have some agency in the process. The experience was positive when their files were retrieved quickly, queues were short, and registration proceeded efficiently. It was, however, exceedingly frustrating when their clinic files were lost, additional payments were required, and system outages occurred as they often had to wait or return home. If they did make it past registration and into the waiting room, sometimes late-arriving patients were served first without explanation, which was also frustrating.

The waiting area presented both opportunities and challenges for support. In general, having to wait at many points during one visit was frustrating. The frustration was alleviated with complimentary porridge, occasional health talks, interactions with other patients, and encouragement from staff. Seeing and talking with other women undergoing cancer care created hope and a sense of belonging. But prolonged waiting time, perceived preferential treatment, reports of bribes, and concerns about missing medicines led to frustration and discouragement. These experiences suggested that frustration was not simply due to lost time, but more to unfairness and a lack of system accountability for causing longer wait times.

> “We felt that certain patients had preferred treatment from providers who were either known to them or received a bribe to allow them to skip the queue and receive treatment quickly while at the clinic.” (PID 016)

#### c) Clinical Examination

Women’s experiences in clinical rooms could similarly reinforce motivation to continue or discontinue engagement with care. Women described feeling welcomed when providers received them warmly, explained results clearly, arranged translation when needed, and communicated difficult results in a caring way. Beginning treatment could itself generate optimism because women felt that action was being taken against the cancer. As one woman explained,

> “I felt good, thinking that now they have started to give me treatment, I feel like I will be well.” (PID 001)

In contrast, meeting unfamiliar or new providers, the logistics of physical examinations, language barriers, treatment side effects, inadequate equipment, and uncertainty about treatment outcomes could produce anxiety and discouragement. Privacy was a key concern for both patients and providers, especially when trainees were present during examinations. One patient explained,

> “You feel some fear to open up and have them look at you…but because you are sick, it forces you just to do as required.” *(PID 008)*

When one patient felt her privacy was not being respected by the trainees, on her way out, she told other women in the waiting room, and some left before even being attended to..

## Laboratory and pharmacy, imaging, and other services

Laboratory testing was valued for providing diagnoses and results, and women were happy to receive medicines when available. However, these services turn stressful when the test costs exceeded their resources, there were long waits due to power outages, their samples or results were lost, the draws were painful, and medicine stock-outs necessitated purchasing medicines elsewhere. One participant described how a power outage could turn a routine laboratory visit into an extended stay:

> *“We sleep in the lab! It forces you to look for a place to sleep and come back the following day.” You come, and you are again given another appointment to come back another day.” (PID 018)*

Imaging was similarly valued because it clarified diagnosis, stage, and treatment plans, but inability to pay, equipment breakdowns, delays, stock-outs, and repeated appointments could produce substantial anxiety and hopelessness.

> *“If you cannot pay, you feel bad because you need the images to get to the doctor, to know how to help you.” (PID 006)*

## Checking out and preparing for the next visit

At checkout, women described relief and gratitude after completing a long day and receiving care. Yet the end of one visit also marked the beginning of planning for the next. When asked what might prevent them from returning, participants identified a lack of money, negative interpersonal experiences, and advice to seek traditional treatment instead of hospital care.

> *“If not money, you might feel offended by someone who is in that place until you say that you will not go back to that place” (PID 012)*

> *“People told me that I would not be healed by coming to the hospital, but [to] go to the traditional healers… When it was not working… I focused on coming to the hospital and got help.” (PID 003)*

Providers additionally identified appointment-scheduling problems, staff-client interactions, and a lengthy care process as challenges to return. Thus, the care journey did not end at checkout; women immediately began considering the financial, interpersonal, and health-system factors that could shape their return.

## Between Days 1 and 2: Research Team Design Synthesis

The experience diagram revealed the precise points during the visit experience where the patient experience broke down. The research team performed affinity clustering on the 55 pain points and, through discussion and consensus-building, identified six pain points that might be remedied through PIF interventions (Table 2). The discussion that led to the identification of those 6 is summarized by the following design insights:

1. Motivation was present at the start of the care journey but declined over time due to financial pressures and repeated system failures. Given that system failures may likely continue, preserving and reinforcing motivation is key to engagement and retention in cervical cancer care.
2. Encouragement and interactions within the clinic environment were described as important to participants’ experiences of care. Participants repeatedly linked a) encouragement for providers and fellow patients, b) welcoming interactions, c) clear communication, d) translation support, and e) receiving treatment to the feeling of being cared for.
3. Challenges in the cervical cancer care journey cut across touchpoints. A disruption in care in one stage could subsequently create additional barriers later in the journey. This suggests that retention should be considered at each touchpoint and as a product of accumulating care experiences.
4. Waiting at the clinic was an experience that holds both opportunities and challenges. Informal patient-to-patient interactions while waiting created opportunities to share and feel less alone, and fostered moments of reciprocal encouragement and social connection. But missing files or being cut in line produced a kind of waiting that was discouraging and thwarted engagement with care.
5. Women valued cervical cancer care services even when accessing them was difficult. For example, the women knew imaging was key in determining the disease, but delays, inability to pay, or machine breakdowns transformed their motivation into hopelessness. Sustaining their engagement with care despite challenges is critical for retention.
6. Generosity existed within substantial financial constraints. Participants expressed willingness to offer mutual support and give what they could despite their own challenges.
7. The end of one visit was the beginning of the next retention decision. Check-out presents an opportunity for the health system to make a long-lasting connection with the patient by learning how the just-ended visit might impact the next visit, seeding hope and motivation, and acknowledging or taking accountability for system failures.

**Table 2:** Initial PIF prototypes

|  | **Pain Point** | **PIF Prototype** | **Prototype Description** | **Fit with PIF** |
| --- | --- | --- | --- | --- |
| 1 | The first oncology clinic visit can be discouraging and unfamiliar especially without adequate support and information. | Peer navigation for new patients. A woman attending her first cervical cancer visit receives orientation, encouragement and support in navigating clinic services. | During the patients first visit, they are offered a peer navigator to walk them through the visit and answer questions. When the patient checks out at the end of the visit, they are asked if they would like to perform this service for another patient sometime in the future. | Peer support reduces anxiety and provides encouragement to remain engaged. |
| 2 | Transportation costs represent the greatest barrier to accessing care, compounded by need to cater for caregiver transport and accommodation costs. | Transport voucher fund | A transportation voucher is offered to selected patients (e.g. clinic nurse selects every 10th patient), who are then asked whether they would like to contribute | A small amount of money can go a long way to ease the cost of transport |
| 3 | Insurance will cover many costs that patients find burdensome but there are challenges to paying the premiums in a lump sum. | Health insurance support fund | When patients cannot pay for cervical cancer care, a gift to cover the cost of insurance is offered. Patients are offered a chance to contribute to the Social Health Assurance (SHA) fund. | Covering the cost of the premiums will encourage patients to take |
| 4 | patients wait at nearly every stage of the care journey, and consistently experience this negatively | Message board in clinic waiting rooms | A message board in the waiting room where patients can write and post encouraging notes. | Encouragement as a bridge during times of waiting |
| 5 | Service integration has benefits for patients' health but adds unexpected cost at the visit where cost is already an issue. | Integrated service gift | A receiving treatment for cervical cancer is offered a test for diabetes or other non-communicable disease. She is told the cost of this test has already been paid for. At her next visit she may pay so that another patient receives the same test for free. | Small sums of money allow patients to access important diagnostic tests and medications for co-morbidities |
| 6 | The visit is long, with much waiting and frustration, in addition to the discomfort and pain of cancer and chemotherapy | Basket of kindness at check out | At checkout patients are told there is a basket of free goods that they may take from if they are in need (e.g. fare to get home, food) They are told they may add to the basket next time, to help another | A small gift acknowledges and eases immediate needs and builds connection with the clinic |

### Day 2: Participant reactions to the initial PIF prototypes

Participants expressed support for all six PIF prototypes while also raising important considerations for their implementation and refinement. We grouped these reactions into two categories: monetary and non-monetary PIF strategies

## Monetary PIF Strategies

These include the transport voucher fund, an integrated service gift, and the health insurance support fund. Participants reported that assistance with health insurance premiums and transport costs was relevant and necessary, expressing that reducing these financial barriers would motivate return for scheduled care. However, concerns about accountability and transparency in fund management were raised, citing reports of corruption in health facilities. Some emphasized the need for formal oversight of the fund by a trusted external body independent of the facility, such as a church or a patient-led self-help group, while others preferred a more collective approach to supporting one another than the more external pay-it-forward model.

> *“Selfishness may lead to corruption from the custodian/lack of integrity. Use existing groups to continue with donations.” (PID 011)*

Participants recognized that the spirit of generosity or a chain of kindness existed; however, it was somewhat diminished by the reality that many people do not have what they need to reliably get to/from, and pay for, everything during a clinic visit, despite their willingness to contribute and support the care of someone else.

> *“Barriers to pay it forward include difficulty returning to the clinic when not receiving treatment and other competing obligations.” (PID 003)*

Some reported that certain health facilities in the region operated an informal kitty or support fund to assist patients in need, which could be leveraged for a PIF initiative. This fund is supported by patients.

> *“Women can be encouraged to donate whatever small amount they have as they view it as a community resource (like the existing kitty).” (PID 008)*

## Non-monetary PIF strategies

This included peer navigation, the message board, and the basket of kindness. These strategies were reported to boost clients’ self-esteem by fostering a sense of belonging and care.

> *“When you leave the clinic, after you meet with peers, you’ll be happy because you will have felt the sense of belonging.” (PID 014)*

Patient navigation at the first visit was reported as an important opportunity to address stigma, provide hope, and motivate patients to return to care. Participants noted that peer navigation was already used in HIV care, providing a similar model that could be adapted for cervical cancer. However, they emphasized that support should be responsive to patients’ readiness for interaction, especially at diagnosis.

> *“It would be helpful if someone showed you around on the first day, but when you are first diagnosed, you may not be in the mood to make a new friend.” (PID 023)*

Participants emphasized the need for continued psychosocial counselling to build confidence and encourage the patient to accept the treatment. The cervical cancer disease process was described as not only financially challenging but also emotionally overwhelming, especially among women who did not receive support from their partners, who are the main decision makers in the household. Participants expressed mixed feelings about male involvement in the cancer care process at the first clinic visit and subsequent visits. Some suggested that involving them through the entire process and also offering psychological support could reduce misunderstandings and relationship strain due to the disease process.

> *“One woman had oozing wounds, and her husband was discouraged and tired of the smell in the home caused by the wounds. The clinic staff spoke to him to help him understand the disease process.” (PID 013)*

> *“Some patients say that their husbands do not believe them when they say they are sick. We also know of cases where a cancer diagnosis has led to separation.” (PID 005)*

Additionally, the long wait patients endure provides an opportunity to connect, learn, and engage by sharing encouraging messages and notes from each other and clinic staff. While the strategy on message boards appealed to many, given that it had been implemented in HIV clinics, writing notes raised concerns about literacy, and patients proposed alternative mechanisms for those with literacy challenges, such as visuals.

## Iterative refinement and feasibility assessment

Following the initial reactions, participants in their small-group discussions refined the prototypes based on their assessments of each prototype’s acceptability and feasibility. The PIF strategies underwent implementation-level refinements while preserving the underlying reciprocal intent.

For the transport voucher fund, the participant acknowledged that not all patients required transport assistance and recommended a needs-based screening prior to invitation, led by social workers and nurses, so that vouchers reach those in need and subsequent donation requests are directed appropriately.

Refinements to the health insurance support fund included oversight by a cancer survivor and a health care provider to enhance transparency and proposed extending the premium gift beyond patients with cervical cancer to other hospital departments, increasing reach while maintaining the donation target to support retention in cervical cancer care.

The integrated service gift refinement included a proposal to accept non-monetary donations (e.g., food and clothing) for sale or distribution in support of the fund. However, operational details were not fully explored and specified.

These refinements were guided by the principles of transparency and accountability. Participants also noted that the components of the three proposed strategies-peer navigation, transport voucher, and health insurance support have been previously implemented in Kenya for other health services, providing locally familiar models that could be adapted for cervical cancer care.

Following the workshop, the research team synthesized participant feedback after refining each prototype. The team then applied the Proctor et al^28^ framework to specify each strategy’s actor, action, action target, temporality, does, implementation outcome, and justification. The resulting PIF strategies are presented in Table 3 below.

**Table 3:**
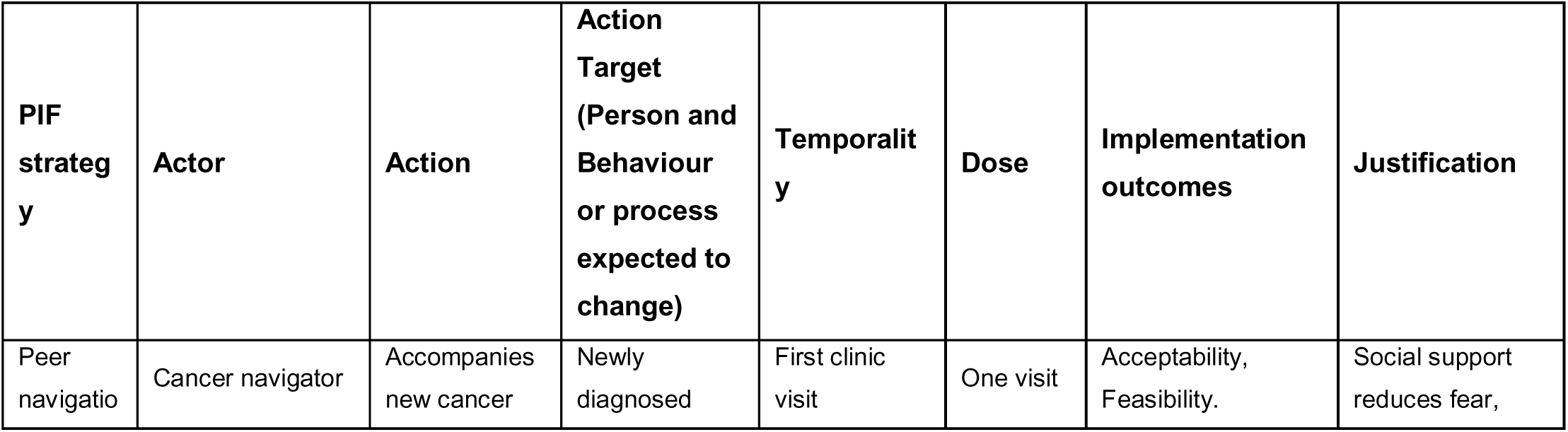
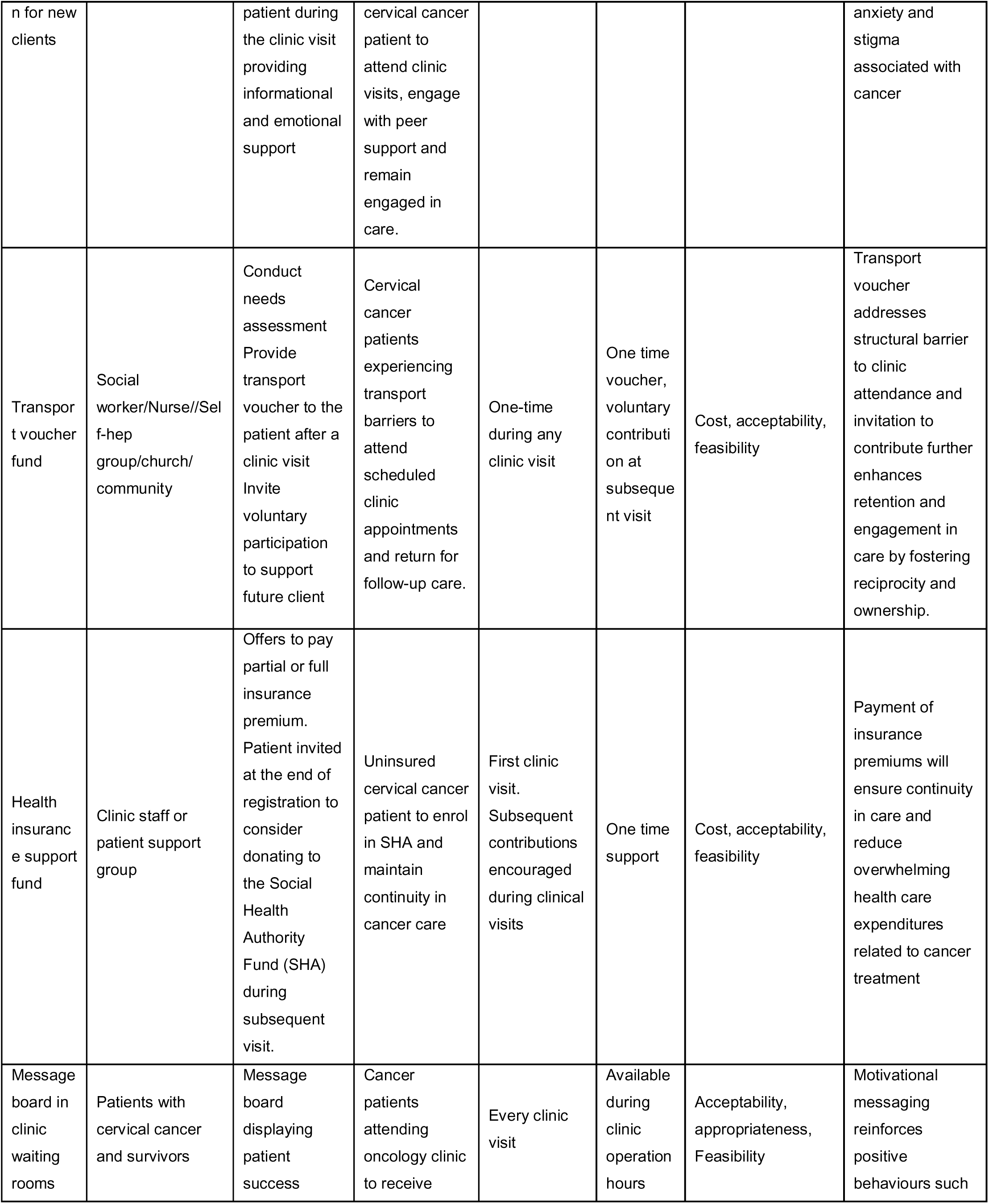
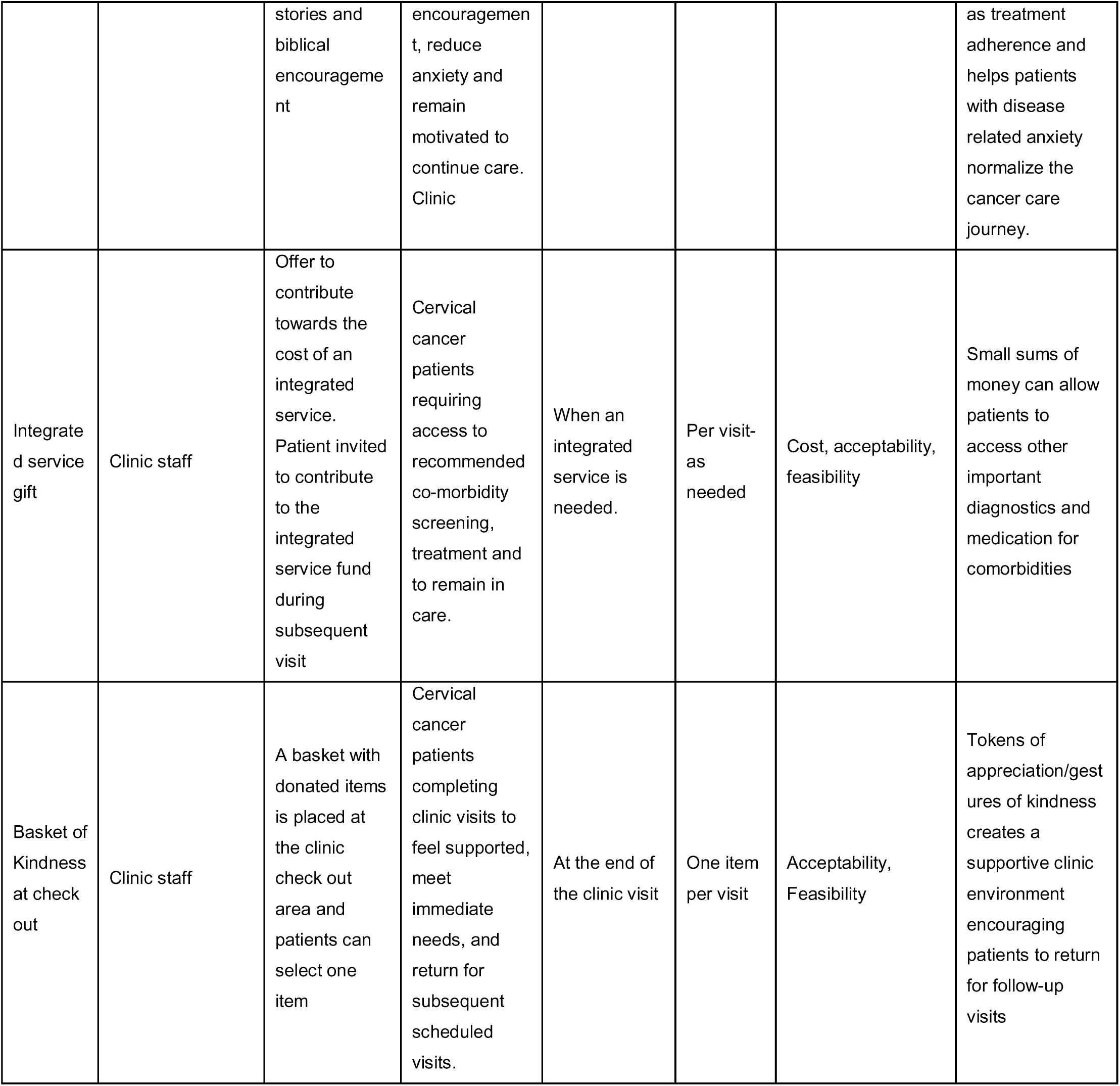
Final co-designed PIF strategies, specified per Proctor et al.

## Discussion

We applied human-centered design (HCD) to co-design PIF strategies for improving retention in cervical cancer care. Unlike conventional qualitative approaches, HCD moves beyond characterizing a problem to collaboratively generating solutions with end users through empathy and structured creativity.^29^ Working with women receiving treatment, survivors, providers, and community representatives, we used experience mapping to generate design insights on where the cervical cancer care journey breaks down for patients and inform six PIF component strategies that might serve to enhance retention. These insights showed that women were often motivated when starting care; however, sustaining that motivation became more challenging as they moved through the journey. The accumulation of financial, logistical, health-system, and treatment-related challenges contributed to frustration, hopelessness, and could undermine continued engagement in care. Positive experiences, on the other hand, could reinforce motivation, highlighting multiple points along the journey where PIF could support continued engagement.

Treatment costs emerged as a significant barrier to engagement with care. Participants described treatment costs as more than a single expense; they included lost income, caregiver costs, and accommodation for women traveling to seek radiotherapy services outside the region.^30^ These financial challenges have been widely reported as a major driver of disparities in cancer survival outcomes. In our setting, these costs are burdensome given the high cost of cervical cancer treatment relative to the average earnings and additional expenses that may not be fully covered by insurance. Importantly, PIF does not require equal financial contributions from all participants. Rather, it provides an opportunity for individuals to contribute voluntarily according to their capacity, including through non-monetary PIF forms of support. Previous PIF research has demonstrated participation across communities with varying income levels, suggesting that even those with low incomes need not be excluded from reciprocal models of care.^31^ In Kenya, where substantial socioeconomic inequalities exist, a flexible PIF model could enable those with greater financial capacity to contribute more while allowing others to participate through other non-monetary forms of support. Such an approach may promote collective participation without placing additional financial expectation on those already experiencing economic hardship.

Beyond financial barriers, participants identified other challenges that could hinder engagement in cervical cancer care, such as extenuating circumstances, discrimination, health system failures, treatment side effects, and fear and anxiety. These barriers are consistent with the literature and not unique to our setting.^32,33^ On the other hand, experience mapping also revealed that supportive interactions could reinforce motivation and enhance care engagement. The end of each visit was also particularly important as the experience there could shape the decision to return. The findings suggest that improving engagement and retention requires not only reducing the financial burden but also strengthening navigation across the cancer care continuum.

The design insights informed PIF opportunities by leveraging the Kenyan spirit of “Harambee”. This included peer navigation, a transport voucher fund, a health insurance fund, an integrated service gift, a basket of kindness at checkout, and a message board in clinic waiting rooms. We have previously shown that peer navigation, messaging, and transport vouchers improve adherence to treatment, viral suppression, and retention in care for people living with HIV.^34–36^ However, these interventions relied on external support, which was difficult to sustain after the studies were completed. PIF could offer an opportunity to integrate them into health systems and community structures while fostering reciprocity and social connection.

Our PIF model differs from prior PIF work in important ways. First, prior PIF studies address discrete, low-cost, preventive services (e.g., a vaccine dose, STI testing).^13,16,37–39^ Cervical cancer care is longitudinal and expensive, and the population asked to give is also the same that bears the financial burden of the disease. Participants described both willingness to contribute alongside difficulty reliably funding their own attendance. Therefore, the sustainability of PIF in oncology will need to be tested in larger studies. In addition, it is unclear whether patients may engage more readily with non-monetary than monetary strategies, given the existing financial burden. Second, prior PIF work included small gifts and brief transactions, without the need for extensive oversight beyond the health facility.

Participants in our study strongly recommended oversight either independent of the facility (e.g., a church or self-help group) or jointly with the facility (a survivor-provider pair). This underscores the importance of transparency and trusted oversight for monetary PIFs.

Drawing on these findings, we hypothesize that PIF operates through a mechanism of social capital activation, transforming patients from recipients of care into participants of a shared moral economy of health (Figure 2).

**Figure 2:**
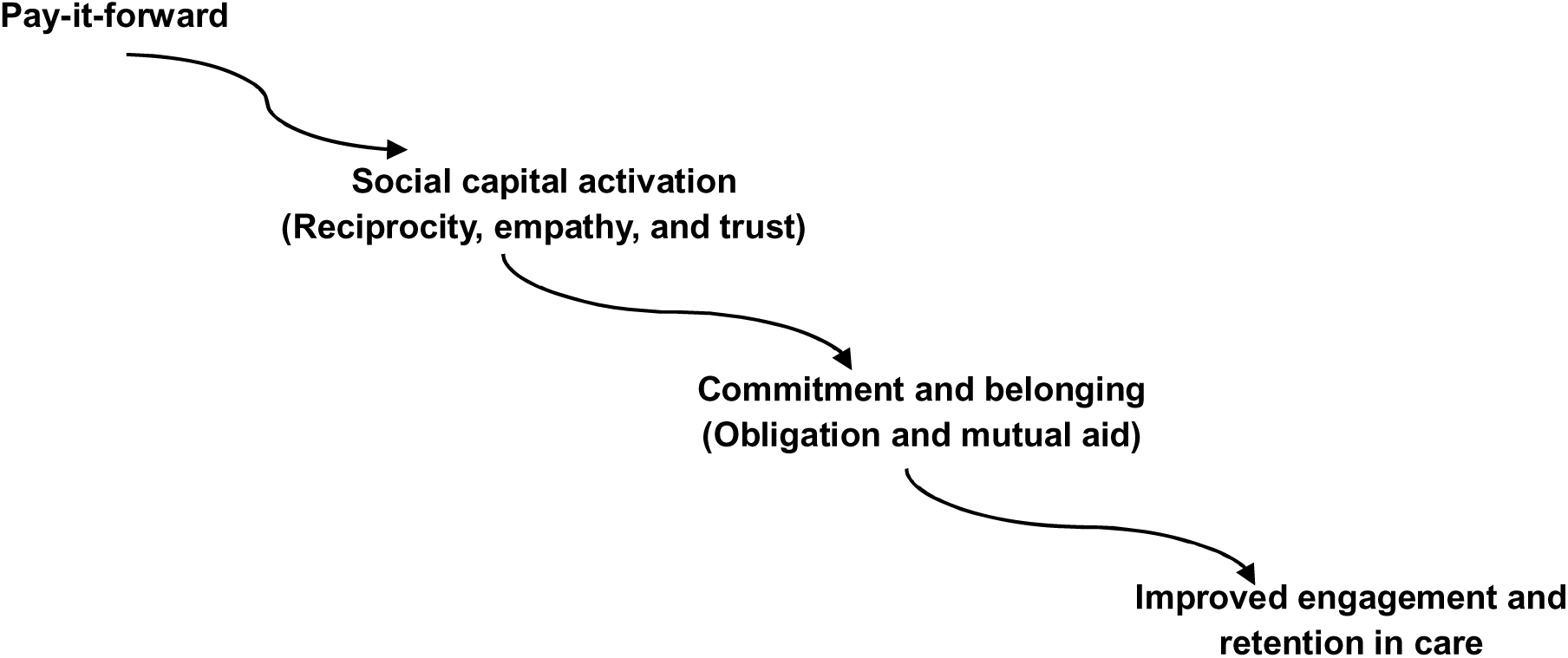
Hypothesized PIF Mechanism

PIF may activate both bonding and bridging social capital.^40^ Bonding social capital emerges through shared patient experiences, especially when patients recognize that others in similar situations contributed to their care, whereas bridging social capital arises when one contributes to an unknown future patient. Each act of giving or supporting another patient creates a relational bridge, forming a system of mutual support that extends beyond the clinic. Through this process, PIF converts individual experiences of care into shared social meaning. The act of giving becomes a signal of belonging and trust, activating bonding (within-group solidarity) and bridging (cross-group empathy) social capital. These social processes generate commitment mechanisms. Therefore, patients stay in care not just because it is medically necessary, but because participation makes them feel socially valued.

Importantly, this mechanism also highlights PIF’s potential to address intersectional barriers that impact care engagement.^10^ For example, women with both cervical cancer and HIV face dual stigma, in addition to financial constraints and health system barriers. PIF may mitigate these challenges by fostering peer connection beyond the cancer clinic encounter.

A key strength of this study is the inclusion of diverse participants, including women receiving cervical cancer treatment, survivors, women living with HIV, providers across different cadres, and community representatives. Experience mapping further helped translate these perspectives into actionable design opportunities.

There were several limitations. First, participating women were all engaged in care. Women lost to follow-up, who may have faced barriers that differ systematically from what we captured, could not be represented. Second, this workshop was conducted at a single site with a sample from a referral hospital, and findings may not be readily transferable to lower-level facilities or other regions. However, cancer care in Kenya is increasingly becoming centralized to large referral hospitals such as our study site. Third, stated willingness to donate may not predict actual donation. However, Kenya, despite being a middle-income country, is ranked the second most generous country in the world^41^and is well known for the national spirit of pulling together, enshrined in the “harambee” motto. Finally, despite emphasis on confidentiality and an open environment, the group format may have limited capturing of all perspectives and experiences.

## Conclusion

Human-centered design offers an opportunity to center end-user voices and create targeted solutions that meet their needs. We identified barriers to retention in cervical cancer care in Kenya and co-designed contextually tailored monetary and non-monetary PIF strategies to address them. Our findings have implications for the development of pro-social health interventions that mobilize social capital and collective action to strengthen health systems. Implementing PIF strategies is not only about delivering an evidence-based intervention but also about mobilizing latent social resources, including empathy, obligation, and mutual aid, that sustain engagement over time. Our co-design approach may also be adaptable to low-resource settings in high-income countries. Future work should focus on piloting these PIF strategies to determine their feasibility, acceptability, and appropriateness, and on establishing whether reciprocal giving can be sustained in this context.

## Data Availability

All data produced are available online at https://figshare.com/s/8a1cf9e6aacb47ec31df

## List of Abbreviations

1. HCD: Human-centered design
2. HIV: Human immunodeficiency virus
3. HPV: Human papillomavirus
4. JOOTRH: Jaramogi Oginga Odinga Teaching and Referral Hospital
5. KEMRI: Kenya Medical Research Institute
6. LFTU: Lost to follow-up
7. LMICs: Low- and middle-income countries
8. PIF: Pay-it-forward
9. SHA: Social Health Authority
10. WHO: World Health Organization
11. UNICEF: United Nations Children’s Fund

## Declarations

### Ethics approval and consent to participate

All participants provided written informed consent. The study was approved by the institutional review boards of the Kenya Medical Research Institute (KEMRI; SERU No. 5210), Jaramogi Oginga Odinga Teaching and Referral Hospital (JOOTRH), and Washington University in St. Louis.

## Consent for publication: N/A

### Availability of data and materials

Data is available at https://figshare.com/s/8a1cf9e6aacb47ec31df

## Authors’ contributions

Conceptualization: TAO, EG, JT. Funding acquisition: TAO.

Scientific oversight and supervision: TAO

Human-centered design methodology: HFA, AT, DM

Workshop design and facilitation: HFA, AT, PKM, LA, JK, BO, DO.

Participant recruitment and data collection: HAF, DM

Acquisition and curation of data: HFA, AT

Qualitative analysis and synthesis: HFA, AT

Development and refinement of PIF strategies: HFA, AT, DM, ET, PKM

Interpretation of findings: HFA, AT, JT

Writing, original draft: HFA

Writing, major revision, and finalization: HFA, TAO, AT, JT

Critical review, editing, and approval of final manuscript: All authors.

## Contributions to Literature

- We propose a hypothesized mechanism through which pay-it-forward may improve engagement and retention in care by activating social capital through reciprocity, empathy, trust, commitment, and belonging.
- We provide a menu of monetary and non-monetary pay-it-forward strategies that can be adapted and tested across different health care settings, offering practical options for addressing financial and psychosocial barriers to retention.
- We extend the pay-it-forward literature beyond one-time, low-cost preventive services by applying the approach to long-term, high-cost cancer care, where sustained engagement presents distinct implementation challenges.

## Acknowledgements

We sincerely thank the women with lived experience of cervical cancer, health care providers, and community representatives who participated in the human-centered design workshop and shared their experiences and perspectives. We also acknowledge the staff of the Kenya Medical Research Institute, Center for Family Health Research and Development (CEFERD), and Jaramogi Oginga Odinga Teaching and Referral Hospital for their support in conducting this study.

